# Who was wearing a mask in 2021? Update on gender-, age-, and location-related differences during the COVID-19 pandemic

**DOI:** 10.1101/2022.01.18.22269479

**Authors:** Michael H. Haischer, Rachel N. Beilfuss, Meggie Rose Hart, Lauren Opielinski, Emma Schmit, David Wrucke, Helena Zhao, Toni D. Uhrich, Sandra K. Hunter

## Abstract

Previous observational work from 2020 demonstrated gender-, age-, and location-related differences in mask-wearing behavior, despite the efficacy and public health messaging that emphasized face coverings in combatting the spread of COVID-19. In 2021, COVID-19 vaccinations and a corresponding change in public health policy became new considerations in deciding personal protective behaviors. To provide an update on mask wearers and resistors approximately one year after our initial study, we observed shoppers (*n* = 6,118) entering retail stores using the same experimental methodology. Approximately 26% of individuals wore a mask. Mask wearing has decreased across demographic groups compared to 2020. Aligning with previous findings, females were ∼1.5x more likely to be observed wearing a mask than males, and the odds of observing a shopper wearing a mask in a suburban or urban area was far greater than at rural stores (∼5.7x and ∼3.3x, respectively). Gender and location are confirmed to be significant and stable factors that impact mask-wearing behavior in the United States during the COVID-19 pandemic. The impact of age on mask wearing was heavily reduced compared to 2020, potentially due to the availability of COVID-19 vaccines and change in mask guidance for vaccinated individuals.

## Introduction

The respiratory virus that causes coronavirus disease 2019 (COVID-19), SARS-CoV-2 [1], is transmitted primarily through the air in small particles that can travel more than six feet [2,3]. Despite the strong public health focus on cleaning and sanitizing touch surfaces [4], evidence suggests that fomites do not appreciably contribute to the spread of the virus [5]. On the other hand, mask wearing and adequate ventilation and filtration of indoor spaces where air is shared by individuals are essential layers of protection to minimize the short-term (e.g., cases, hospitalizations, deaths) and long-term (e.g., “long COVID”) impact on public health [2]. Controversy on the issue of face coverings has continued throughout the COVID-19 pandemic, largely politicized and in rare cases even leading to violence [6-9]. Even so, scientific experts in a variety of fields agree that masks are a safe and effective way of reducing airborne transmission of SARS-CoV-2 [2,10-12]. Similarly, though responses vary considerably across political party lines, polling from August 2021 suggested most Americans favor mandatory masking in public spaces [13]. Epidemiological studies have shown mask wearing to be associated with a reduction in cases, hospitalizations, and deaths [14-16]. Schools with mask policies demonstrate a better ability to limit spread among children [17], and though the acute illness may be mild in most pediatric cases [18], an alarming percentage of individuals of all ages suffer long-term health complications [19,20]. The unknown lasting public health implications of the pandemic and the chronic effects of post-acute COVID-19 syndrome (i.e., “long COVID”) within individuals highlight the importance of considering future public health burden in adopting personal protective measures. While the effectiveness of masks largely depends on material and fit, face coverings may confer protection to both the wearer and individuals in spaces where air is shared [21-23]. To that end, health messaging that encouraged mask wearing gained strength in the United States summer 2020 (June - August), with many public entities and private companies eventually requiring masks for all except young children [24,25].

To study the demographics of mask wearers during summer months of 2020 in the USA, we conducted an observational study and systematically recorded the behavior of individuals entering retail stores [26]. We found that in June 2020, women (gender expression), older adults (estimated age), and those in urban area stores (location) wore masks at the highest levels [26]. Importantly, follow-up observations collected after mask mandates were announced or implemented (July-August 2020) showed over 90% of individuals were wearing masks. Thus, public health policies addressing masks have great impact on the personal protective behaviors of retail shoppers.

In early 2021, COVID-19 vaccinations became widely available for individuals over the age of 16 under an emergency use authorization from the United States Food and Drug Administration [27]. However, adoption of vaccines among the public was moving slower than United States officials hoped [28]. To help encourage vaccinations, the Centers for Disease Control and Prevention changed their policy on May 13^th^, 2021, to state that vaccinated individuals do not need to wear masks [29]. This change in policy and the increased prevalence of the Delta variant of the SARS-CoV-2 virus, known to be more transmissible and potentially more deadly than earlier strains [30,31], introduced new considerations into personal protective behaviors. Thus, based on the new developments in public policy resulting from the availability of vaccines, the aim of this study was to update mask use by gender expression, estimated age, and location, approximately one year after the study conducted in June 2020 [26]. Due to the availability of vaccines and corresponding change in public health messaging, we *hypothesized* that mask use would be reduced in all demographic groups compared to 2020. Further, we *hypothesized* that mask use among females would be greater than males across all age groups and locations, aligning with our previous study and others demonstrating that a greater percentage of females adopt personal protective behaviors [32]. Individuals aged 12-15 years were not able to get the vaccine until May 10^th^, 2021 [33] and adults, especially those in advanced age, were strongly encouraged to get vaccinated. While data suggests rural counties receive COVID-19 vaccinations at lower rates than urban counties [34], rural shoppers wore masks at lower rates in our initial study compared to urban and suburban store-goers [26]. Consequently, we *hypothesized* that mask use would be approximately equal across age groups but would still be less common in rural than in urban or suburban areas.

## Materials and Methods

Our primary aim was addressed by observing mask wearing in the same experimental manner at the same retail locations in southeastern Wisconsin as the previous study [26]. The center point used to define location was in the city of Milwaukee (United States Postal Service main office; <u>345 W St. Paul Ave, Milwaukee, WI, USA)</u>. Thus, stores were placed into the same urban (<6.1 km from city center; *n* = 15, 3.2 ± 1.8 km to city center), suburban (11.5-32.1 km; *n* = 13, 20.5 ± 7.2 km), and rural (>36.9 km; *n* = 9, 55.8 ± 21.4 km) store groups. Visits to stores (*n* = 37) occurred between 9:00 am and 8:00 pm (June 8^th^–28^th^, 2021) and lasted approximately 44 minutes on average. Shoppers were not aware they were being observed and children under the estimated age of two were not recorded. Age categories were defined as young = 2-30 years old, middle age = 30-65 years old, older: >65 years old, aligning with the increases in stratified death rates [35]. Summary observation sheets were crosschecked by other observers and all procedures involved public observation or information and did not require review by an Institutional Review Board.

The impact of gender, age, location, and their interactions on mask wearing was determined via multiple logistic regression analysis. Only individuals that could be grouped into a dichotomous outcome (mask/no mask) were considered. Thus, individuals who were wearing their mask or face covering improperly (not over nose and mouth) were recorded but excluded from further analyses (*n* = 52). Observations of the remaining 6118 individuals were dummy coded for gender expression, age, and location independent variables before being entered into a backward elimination regression. Limit for variable removal and test classification cutoff were set at 0.025 and 0.5, respectively. Adjusted odds ratios (aOR) are expressed with respect to reference groups (gender: male, age: young, location: rural) with 95% confidence intervals and significance was determined at *p* < 0.05. All analyses were performed with either Microsoft Excel (Microsoft, Redmond, WA, USA) or IBM Statistical Package for Social Sciences version 26 (IBM, Armonk, NY, USA).

## Results and Discussion

Over 37 observational visits to retail stores, and 6118 individuals were recorded entering retail stores. They were grouped into a dichotomous outcome (mask/no mask). Overall, 25.7% of individuals were wearing a mask or face covering over their nose and mouth (Fig 1). The trend across gender, age, and location generally aligned with our hypotheses and only young females in rural areas exhibited a positive change in observed odds of mask wearing compared to 2020 (Table 1). Females wore masks 7.6% more than males (28.8% vs. 21.2%; Fig 2A). Additionally, masks were observed at similar percentages across all age groups (young: 27.2%; middle age: 23.7%; older: 27.0%) (Fig 2B). Mask wearing was more in urban (35.9%) than suburban (25.8%) locations and was lowest at stores in rural areas (9.2%) (Fig 2C).

**Table 1.** Observed odds 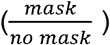 of wearing a mask by age and location* (shown with change in observed odds from Haischer et al., 2020 [26]). Only young females in rural areas showed increased odds of wearing a mask when compared to results from Haischer et al., 2020.

|  | *Young* |  | *Middle Age* |  | *Older* |  |
| --- | --- | --- | --- | --- | --- | --- |
|  | Female | Male | Female | Male | Female | Male |
| *Urban* | 0.545<br>(-0.334) | 0.372<br>(-0.267) | 0.926<br>(-0.096) | 0.443<br>(-0.420) | 1.108<br>(-1.007) | 0.661<br>(-0.388) |
| *Suburban* | 0.442<br>(-0.444) | 0.350<br>(-0.217) | 0.372<br>(-0.885) | 0.203<br>(-0.560) | 0.385<br>(-1.728) | 0.267<br>(-1.201) |
| *Rural* | 0.162<br>(+0.086) | 0.090<br>(-0.027) | 0.162<br>(-0.125) | 0.052<br>(-0.120) | 0.231<br>(-0.863) | 0.175<br>(-0.425) |

\* Odds > 1 show odds of wearing a mask are greater than not wearing a mask

**Fig 1.**
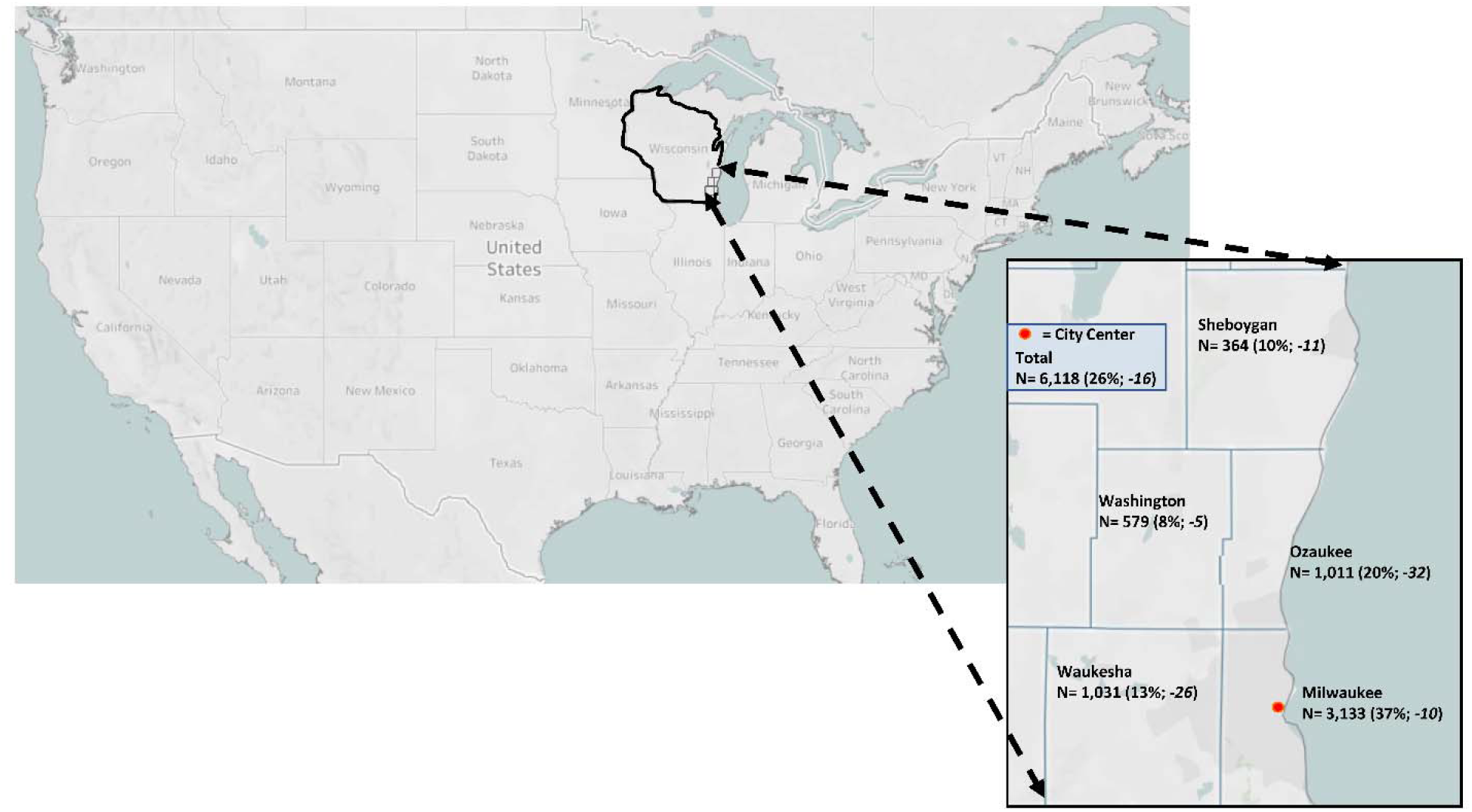
Update on mask-wearing percentages by Wisconsin county. Data shown indicates the number of observations collected and percentage of people wearing a mask (vs. no mask) in each county where retail stores were visited between June 8^th^ and June 28^th^, 2021. Italicized data indicates percentage change in mask wearing from Haischer et al., 2020 [26]. Created in Tableau Public 2020.2.1 (Tableau Software, LLC., Seattle, WA, USA).

**Fig 2.**
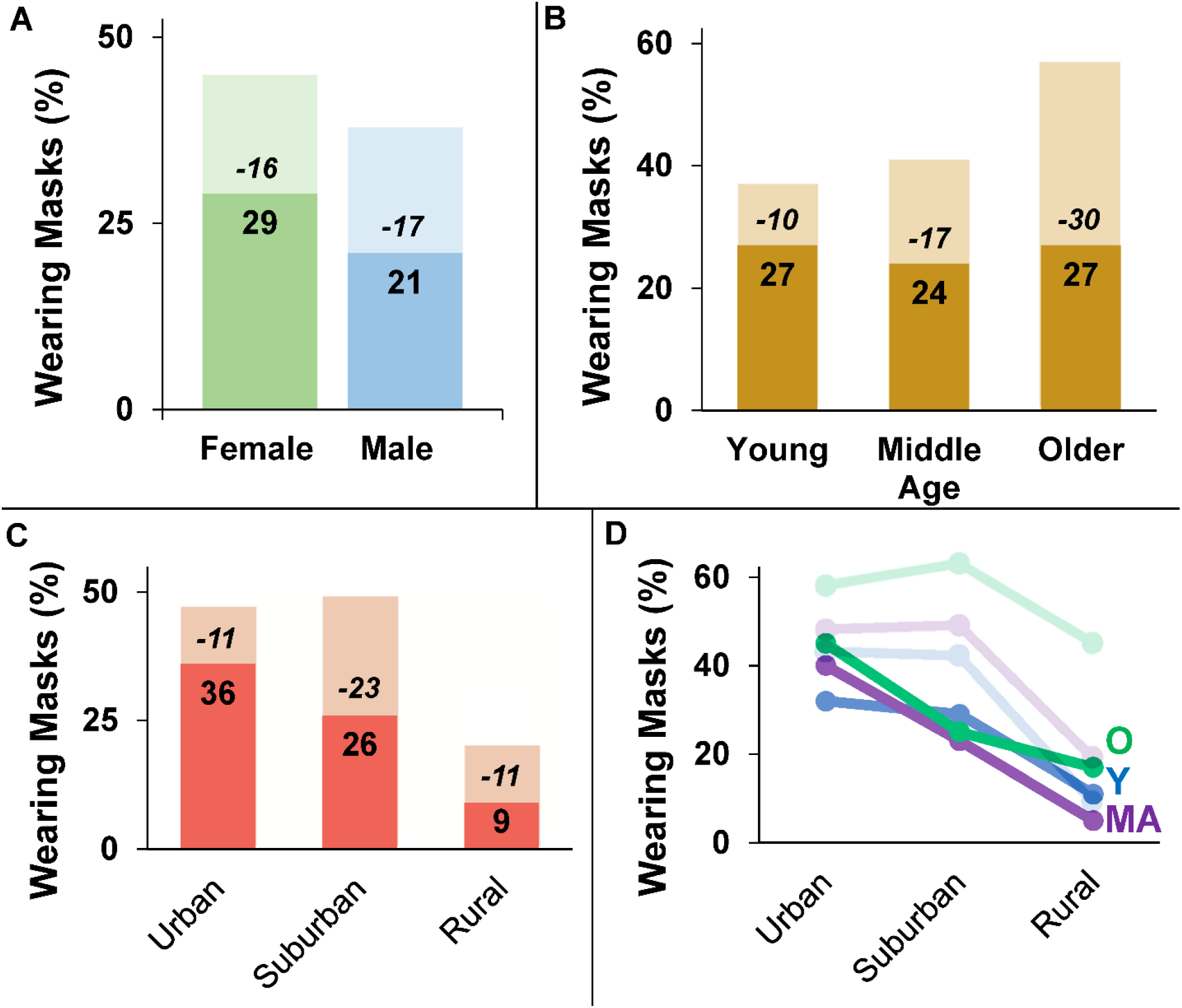
Update on mask-wearing percentages across gender, age, and location. Data indicate percentage of individuals wearing a mask (vs. no mask) between June 8^th^ and June 28^th^, 2021, and percentage change from Haischer et al., 2020 [26] (Darker bars: current study; lighter bars: results from Haischer et al., 2020). **A**. Females still wear masks more than males, but both genders wear masks less than in 2020. **B**. Mask wearing behavior is similar across age groups, unlike in Summer 2020. **C**. Mask-wearing is observed less across all locations and still drops off considerably at rural stores. **D**. Mask wearing plotted by location shows that mask-wearing behavior of all age groups is impacted similarly across geographic areas (older: O; middle-age: MA; young: Y; darker lines: current study; lighter lines: results from Haischer et al., 2020).

Results of the logistic regression analysis indicated that females were significantly more likely to be observed wearing a mask compared to males (aOR = 1.548, 95% CI = 1.369-1.740, p < 0.001) (Fig 3). Additionally, odds of mask wearing were slightly greater for older adults compared to younger individuals (aOR = 1.236, 95% CI = 1.034-1.477, p = 0.020), but did not differ between middle age and younger shoppers (p = 0.449). Coinciding with the low rate of mask wearing at rural stores, the odds of observing a mask wearer was much greater in urban (aOR = 5.712, 95% CI = 4.616-7.086, p < 0.001) and suburban areas (aOR = 3.342, 95% CI = 2.711-4.120, p < 0.001). Unlike the previous study which showed a significant age-location interaction effect [26], the impact of age on mask-wearing behavior was generally consistent across locations (Fig 2D).

**Fig 3.**
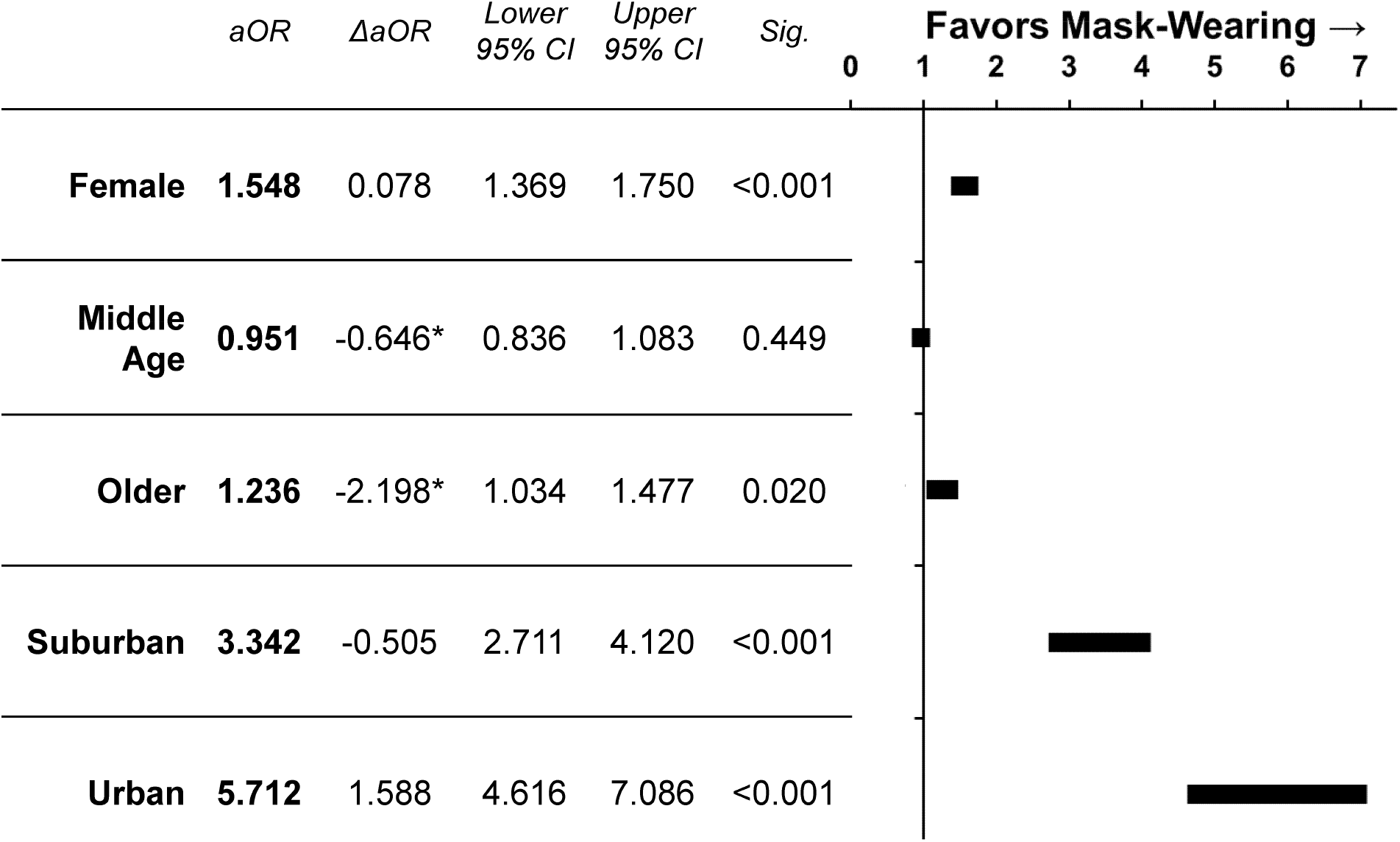
Odds of wearing a mask. Adjusted odds ratios (aOR) and 95% confidence interval plots of mask usage for gender expression, age, and location from June 8^th^ to June 28^th^, 2021 and change in odds ratios (ΔaOR) from Haischer et al., 2020 [26]. Odds ratios are expressed in relation to reference groups (gender: male; age: young; location: rural). *Confidence intervals of aOR do not overlap with Haischer et al., 2020.

When comparing the results of the current study to those from the initial study performed in 2020 [26], there are several notable differences. Overall, mask wearing decreased by about 16% in a year (41.5% in 2020, 25.7% in current study), with similar declines for both males and females (Fig 2A). Thus, despite both genders exhibiting reductions in masking compared to 2020, the odds of observing a female wearing a mask are stable at ∼1.5 times that of males. The explanation for this finding is likely multifactorial but could be related to the perception that masks are seen as emasculating among males in the US [36]. Other factors that may facilitate gender differences in mask-wearing behavior include differences in caregiving responsibilities [37] and preexisting gender inequalities that were exposed due to the pandemic [38].

In examining age groups, mask-wearing rates were markedly different in 2020 but a greater reduction among older and middle-aged adults resulted in similar rates across age groups in 2021 (∼25%, Fig 2B). As a result of this differential reduction in masking, the odds of older and middle-aged individuals being observed wearing a mask compared to younger individuals was markedly changed compared to 2020 (older ΔaOR = -2.198; middle age ΔaOR = -0.646). Consequently, in 2021 there was only slightly greater odds of observing an older adult masking compared to a younger individual and no longer a difference in behavior between middle age and younger individuals. A possible explanation of the greater percentage of older adults wearing masks in 2020 was the higher risk for more severe outcomes from COVID-19 [35]. Consequently, as soon as the emergency use authorization was granted, older individuals were prioritized for COVID-19 vaccines [27,39]. Thus, the change in public health messaging stating masks were not necessary for the vaccinated [29] may be a primary reason for the reduction in mask wearing, especially among older adults.

As in 2020, when considering store location, mask wearing was observed much less frequently in rural compared to urban or suburban areas. However, while mask wearing declined similarly in urban and rural areas (∼11%), the behavior was more heavily reduced in suburban locations (∼23%). As a result, the odds of observing a mask wearer compared to 2020 were increased in urban areas (ΔaOR = 1.588) but reduced in suburban areas (ΔaOR = -0.505). Even so, overlapping confidence intervals of the odds ratios calculated in 2020 and 2021 for these areas suggest the changes are not appreciably different.

Importantly, our overarching findings confirm that many individuals still resist masks despite public health guidance to wear a mask in public locations such as retail stores. As of June 27^th^, 2021, only 49.3% of Wisconsin residents over the age of 12 years were fully vaccinated [40]. Consequently, if there was compliance with public health recommendations, we should have observed approximately half of individuals wearing masks. The difference between overall vaccination rate in Wisconsin and mask wearing in the current study however was ∼24%. Despite the limitations of estimating gender and age, and the availability of vaccines to individuals over 12 years, it can be roughly estimated that up to 25% of shoppers observed during data collection were not vaccinated and were also not wearing a mask. Even asymptomatic individuals may transmit the virus to other people [41-43] and unvaccinated persons likely transmit at a greater rate than vaccinated individuals [44-45]. Taken together with our results, the potentially large number of unvaccinated and unmasked persons in retail stores could have a substantial impact in prolonging the pandemic.

## Conclusions

Despite the efficacy of mask wearing as a personal protective intervention during the COVID-19 pandemic, only 26% of over 6100 individuals entering retail stores in Wisconsin were observed to be wearing a mask. This was a 16% reduction from one year before (2020) in the same locations [26]. Males and shoppers in rural stores wore masks significantly less than females and store-goers at suburban and urban locations, respectively, aligning with findings from the previous year [26]. The impact of age on mask-wearing was considerably diminished compared to 2020, in that only slightly greater odds of mask wearing were observed for older adults in reference to younger individuals. The widespread availability of COVID-19 vaccines and corresponding change in health guidance on masking may be viewed as a possible explanation for this differential result over the year. Considering vaccination rates in the area, we estimated an alarming percentage of shoppers were likely unvaccinated and also not wearing a mask (up to 25%). Our updated findings will help individuals to continue to assess the risks of retail shopping during the COVID-19 pandemic. Key stakeholders may also benefit from considering these results when devising public health policies related to face masks in the current and future pandemics.

## Data Availability

All data produced in the present study are available upon reasonable request to the authors.

